# Carbon dioxide, COVID-19 and the importance of restaurant ventilation: a case study from Spain approaching Christmas 2021

**DOI:** 10.1101/2021.12.17.21267987

**Authors:** Teresa Moreno, Wes Gibbons

## Abstract

Restaurants present an especial challenge in the prevention of the spread of COVID-19 via exhalatory bioaerosols because customers are unprotected by facemasks while eating, so that ventilation protocols in such establishments become particularly important. However, despite the fact that this pandemic airborne disease has been with us for two full years, many restaurants are still not successfully prioritising air renovation as a key tool for reducing infection risk. We demonstrate this in the run-up to the 2021 Christmas celebrations by reporting on CO_2_ concentration data obtained from a hotel breakfast room and restaurants during the 5-day Spanish holiday period of 4^th^-8^th^ December. In the case of the breakfast room, inadequate ventilation resulted in average CO_2_ levels ranging from 868 to 1237ppm on five consecutive days, with the highest levels coinciding with highest occupancy numbers. Inside the five restaurants, three of these were well ventilated, maintaining stable average CO_2_ concentrations below 700ppm. In contrast, two restaurants failed to keep average CO_2_ levels below 1000ppm, despite sporadic, but ineffective, attempts by one of them to ventilate the establishment. More effort needs to be made to foster in both restaurant managers and the general public an improved awareness of the value of CO_2_ concentrations as an infection risk proxy and the relevance of ventilation issues to the propagation of respiratory diseases.

## Introduction

The concentration of CO_2_ in each human exhalation is around 100 times higher than that in outside ambient air. Thus, in a working restaurant CO_2_ levels will quickly rise unless the indoor air is renovated by introducing outdoor air. The concentration of CO_2_ in a public eating area at any given time therefore provides an indication of how successfully the ventilation is managing to refresh the air, that is to say, how much a customer is likely to be breathing air that has passed through the lungs of other people sharing the same space. As such, the airborne infection risk of a given respiratory pathogen would be expected to increase with increasing concentrations of exhaled CO_2_ in a given restaurant, as well as being influenced by the behaviour of the occupants.

COVID-19 is known to be capable of airborne transmission within indoor environments and this method of propagation is fuelling the ongoing pandemic (e.g. Bazant and Bush, 2021; Buonnano et al., 2020a; Greenhalgh et al., 2021; Miller et al., 2021; Morawska and Milton, 2020; Peng et al., 2021; Shen et al., 2020; Wang et al., 2021; Zhang et al., 2021). The dominant mechanism for such transmission is widely considered to involve infected individuals exhaling viable virus-bearing aerosols. These exhalatory “viraerosols” (Moreno and Gibbons, 2021) can be released in great numbers not only by coughing, sneezing and normal vocalisation, but also simply by passive tidal breathing (e.g. Almstrand et al., 2010; Bake et al., 2019; Haslbeck et al., 2010; Johnson and Morawska, 2009; Schwarz et al., 2010). Once released into the indoor atmosphere, the potentially infective respiratory pathogens will be carried away from the diseased individual in a buoyant turbulent cloud of gas and particles (Boubouiba, 2021; Jones and Bross, 2015; Lv et al., 2021; Randall et al., 2021). Thus, to minimise respiratory infection risk in public indoor spaces the obvious recommendation is to wear a well-fitted, high quality facemask, and combine this with efficient ventilation that ensures the constant introduction of fresh air from outside.

The use of facemasks is however not practicable in restaurant settings, so that ventilation protocols in such establishments become especially important. It is fully two years since the emergence of SARS-CoV-2, and there has been a clear call from some in the scientific community that a revolution is needed in the way we think about indoor air quality (e.g. Melikov, 2020; Morawska et al., 2021). Nevertheless, the message that air renovation by ventilation is likely to be of vital importance to reduce the spread of COVID-19 has still not hit home in many public eating places. We demonstrate this by reporting on data collected during the busy nationwide early December Spanish double bank holiday (6^th^ and 8^th^) which is celebrated throughout the country and presents the threat of boosting a viral superspreading event linked to Christmas celebrations during the rest of the month.

In 2021 the early December holiday period in Spain centred on a Monday (6^th^) and Wednesday (8^th^), so many people chose to take a longer break that included the previous weekend. Hotels and restaurants were exceptionally busy, despite concerns relating to the ongoing spread of the highly infectious omicron SARS-CoV-2 viral variant. In this study we collected CO_2_ data during the 5-day holiday period (Saturday 4^th^ to Wednesday 8^th^) from (1) a hotel breakfast room, and (2) whilst eating meals at five different restaurants. In order to ensure we were recording real-life conditions, we did not ask permission to measure CO_2_ in any of these establishments and therefore provide no location details. CO_2_, RH, temperature and PM_2.5_ measurements were obtained at around 50cm above floor level using an IQAir (Air Visual Pro, https://www.iqair.com/en/air-quality-monitors/airvisual-pro) monitor, which measures CO_2_ concentrations between 400-10,000ppm every 10 seconds. The system operates portably for 4 hours using a lithium battery. The person responsible for collecting the CO_2_ measurements in all establishments tested negative for COVID-19 on the 12^th^ December.

## The hotel breakfast room

Table 1 and Figure 1 summarise the data from the hotel breakfast room. This was a large room on the first floor, connected to the rest of the hotel by a door leading to a staircase down to the hotel ground floor (not directly to outside air). This door was kept fully open throughout the breakfast period, but all six windows were kept closed. There was no CO_2_ meter in the breakfast room, although one was operating in the ground floor bar below. All CO_2_ measurements for this study were taken during the same breakfast time-slot (10.00-10.45 am) and from a similar part of the restaurant and at a height around 50cm above the floor. Seating protocols ensured that tables were separated by >1m and they were required to wear masks when helping themselves to the food buffet. Most guests wore correctly fitted masks when standing at the buffet, although there were a few notable exceptions.

**Table 1.** Hotel breakfast room

|  | CO<sub>2</sub> (ppm) |  | RH (%) |  | T (°C) |  | PM<sub>2.5</sub> (µg/m<sup>3</sup>) |  |
| --- | --- | --- | --- | --- | --- | --- | --- | --- |
|  | Mean | Max | Mean | Max | Mean | Max | Mean | Max |
| **Day 1** | 868 | 919 | 56 | 57 | 23,8 | 24,5 | 4 | 7 |
| **Day 2** | 1237 | 1342 | 54 | 56 | 23,5 | 24,1 | 15 | 65 |
| **Day 3** | 1140 | 1180 | 54 | 56 | 24,3 | 25,1 | 6 | 10 |
| **Day 4** | 948 | 1004 | 59 | 62 | 24,9 | 25,2 | 3 | 5 |
| **Day 5** | 955 | 997 | 48 | 48 | 24,1 | 24,2 | 10 | 13 |

**Figure 1:**
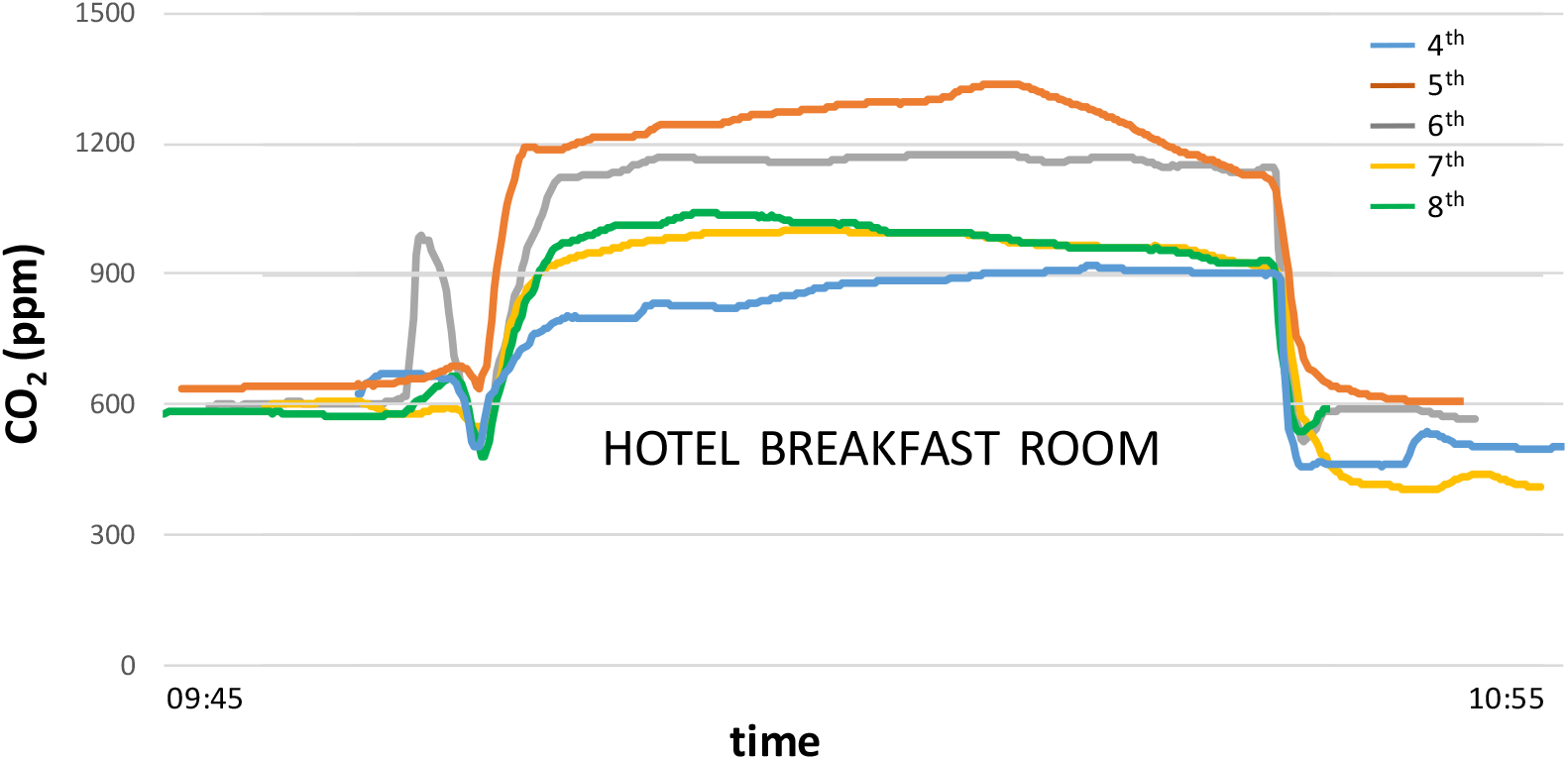
CO_2_ concentration curves for hotel breakfast periods on the five days 4^th^-8^th^ December 2021. The curves record initial hotel room conditions (600-650ppm) interrupted by a brief trough marking a short outdoor walk to the breakfast room inside which CO_2_ levels rose steeply, especially on the 5^th^ (Day 2 on Table 1) when the room was fully occupied. The pre-breakfast transient peak on the 6^th^ records entry into the hotel bar area. See text for details.

The lowest CO_2_ concentrations in the hotel breakfast room (average 868ppm: Table 1) were measured at the beginning of the holiday period (4^th^), before hotel occupancy numbers reached their peak. During this first breakfast CO_2_ levels climbed slowly to a maximum value of 919ppm (Table 1). In contrast, the following day (5^th^), after high hotel occupancy levels during the Saturday night, the breakfast room was exceptionally busy (CO_2_ average 1237ppm). Under these conditions CO_2_ concentrations at 10.00 were already approaching 1200ppm and then climbed to a peak of 1342ppm in the fully occupied room before slowly declining as people began to finish their breakfast and leave the establishment (Figure 1). On the morning of the bank holiday on the 6^th^ December CO_2_ levels were again high (Day 3 average 1140ppm: Table 1), falling to around 950ppm average on the remaining two days (Days 4 and 5: Figure 1). The atmospheric stability of this indoor setting over the five mornings was reflected by minimal variation in both temperature and RH. Apart from the exceptionally busy Day 2 (marked by much customer and staff movement and occasional toast burning events), average PM_2.5_ levels also showed little variation on a given morning (varying by only 2-4 µg/m^3^: Table 1). With the windows closed and the internal entry door left open, the dominant control on CO_2_ concentrations appears to have been the number of guests attending breakfast. On the morning of the second day (5^th^), the headwaiter was made aware of the fact that the ventilation could be better, and agreed with this observation. However, nothing obvious was changed on subsequent mornings: in particular all windows remained closed.

## The restaurants

Five restaurants were chosen to sample contrasting atmospheric scenarios (Table 2 and Figure 2). The data collected indicate that three of these restaurants (A, B, C: Figure 2) were well ventilated, maintaining stable CO_2_ concentrations which averaged below 700ppm and never reached maxima of 1000ppm (Table 2), despite the fact that each establishment was different in character. Restaurant A was small, with few (<20) customers and with the front door continuously held slightly open it registered CO_2_ average levels of 665ppm, although combined with relatively high PM_2.5_ concentrations attributed to cooking emissions. Restaurant B was a narrow bar restaurant (<20 clients indoors), with tables separated by screens, the front door fully open, and showed similar air quality conditions to Restaurant A (Table 2). Restaurant C was a large, busy open-plan converted market building with multiple food outlets and two floors (the monitor recorded on the upper floor) and a prominent new ceiling HVAC system. Despite a multitude of customers and staff housed in this establishment, its large size combined with the HVAC system successfully kept average CO_2_ levels at 684ppm and average PM_2.5_ concentrations at 9µg/m^3^ (Table 2). Despite the considerable differences between these three establishments, all of them thus managed to maintain stable, relatively low CO_2_ levels, punctuated only occasionally by transient peaks due to people clustering near the monitor.

**Table 2.** Restaurants

|  | CO<sub>2</sub> (ppm) |  | RH (%) |  | T (°C) |  | PM<sub>2.5</sub> (µg/m<sup>3</sup>) |  |
| --- | --- | --- | --- | --- | --- | --- | --- | --- |
|  | Mean | Max | Mean | Max | Mean | Max | Mean | Max |
| **A** | 665 | 860 | 55 | 57 | 20,0 | 20,8 | 25 | 59 |
| **B** | 651 | 924 | 60 | 71 | 20,7 | 21,7 | 11 | 38 |
| **C** | 684 | 754 | 47 | 55 | 23,6 | 24,0 | 9 | 26 |
| **D** | 1029 | 1310 | 56 | 58 | 21,9 | 22,4 | 45 | 94 |
| **E** | 1014 | 1116 | 60 | 62 | 21,3 | 21,6 | 16 | 25 |

**Figure 2:**
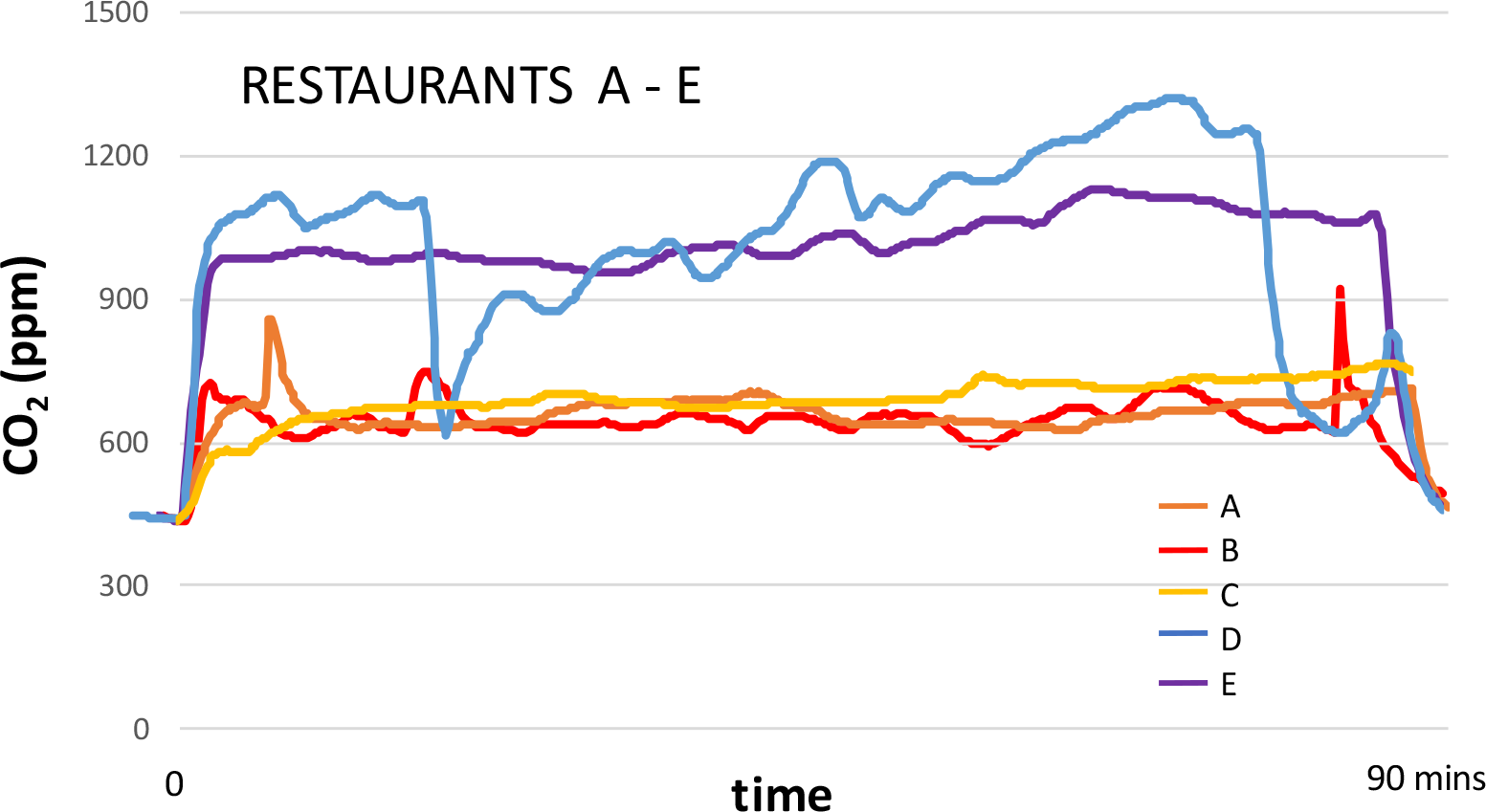
CO_2_ concentration curves for the five restaurants sampled during the study. A, B and C record good, generally stable ventilation conditions, with average CO_2_ <700ppm, in contrast to Restaurants D and E. Whereas Restaurant D made sporadic efforts to ventilate the establishment by opening the front door (marked by two precipitate falls towards 600ppm), Restaurant E kept doors and windows closed. See text for details.

In contrast, two of the restaurants failed to maintain average CO_2_ concentrations below 1000ppm during the meal (D and E: Table 2). In the case of Restaurant D, this was a medium-sized establishment which was fully occupied for much of the time, although maintaining correct adherence to table separation rules. The restaurant owner was aware of the need for ventilation, which she attempted to achieve by fully opening the front door from time to time, allowing a blast of cold air into the room that was not appreciated by some guests. Carbon dioxide concentration data for Restaurant D are shown on Figure 2 and show levels of around 1050ppm on entry into the establishment, with slight declines in the curve recording the arrival of guests briefly opening the front door. Ventilating the restaurant suddenly by fully opening the front door caused CO_2_ levels to drop immediately to 600ppm (Figure 2), but after this levels rose rapidly, interrupted by further guest arrivals, to peak at 1310ppm. A repeated brief ventilation episode suddenly reduced CO_2_ concentrations once again to 600ppm before beginning another cycle of rising levels (Figure 2). Another notable characteristic of this restaurant was its high levels of average and maximum PM_2.5_ (Table 2).

The other establishment failing to ensure minimum levels of CO_2_ was smaller (Restaurant E) and, although much less busy (<20 clients), had chosen to keep the front door firmly closed. This scenario generated stable but high levels of CO_2_ which were nearly 1000ppm upon entry and subsequently rose gradually to peak at 1116ppm (Figure 2; Table 2).

## Discussion and conclusion

The risk of COVID-19 infection in an indoor space will depend on several factors, and modelling calculations based on the mid-20^th^ century works by William Wells, Richard Riley and their colleagues have been applied to the problem by several authors (e.g. Buonanno et al., 2020b; Buonanno and Stabile in Moreno et al., 2021; Bazant and Bush, 2021; Peng and Jimenez, 2021; Peng et al., 2021). Key influencing factors include the number, infectiousness and behaviour of diseased occupants, the room size and air renovation rates, the overall occupancy and duration of exposure, the temperature, relative humidity and the evaporation rate of viraerosols of different sizes (Kin and Marr, 2019). In this context, CO_2_ concentrations offer a proxy for the infection risk although, as emphasised by Peng and Jimenez (2021), this risk varies greatly depending on the exact nature of the indoor setting.

With respect to restaurants, each establishment has its own transiently varying atmospheric microenvironment that needs to be managed, and this dynamic adds local complexity to the problem of ventilation control. To this challenge may be added the vagaries of customer behaviour (including complaints regarding the entry of cold air), concern for increased heating costs and, as we observed in one establishment, the possibility of grossly inaccurate CO_2_ monitors on display. Despite these difficulties, however, it is clear that some establishments are being much more successful than others at maintaining low CO_2_ concentrations, and this is likely to impact on health effects. To take a hypothetical example from our study locations, an asymptomatic but virally loaded COVID-19-positive family conversing enthusiastically over an early breakfast on the 5^th^ December could have left a turbulent cloud of SARS-CoV-2-bearing aerosols dispersing slowly through the poorly ventilated room. Successive waves of breakfast guests arriving according to their pre-planned 45-minute timeslot that morning, as CO_2_ levels rose above 1200ppm, could have been exposed to airborne viruses previously exhaled by the family and thus been potentially vulnerable to infection. If the same family had later dined at Restaurant C, their exhalatory pathogens would have been more quickly dispersed and diluted within a better ventilated atmosphere averaging 684ppm CO_2_, and they would thus have presented less of a potential threat to other customers sharing the same space.

The short and straightforward nature of this study was designed to be equal to the simplicity of the message it contains: in the run-up to our third COVID-19 Christmas some restaurant managers are still not successfully prioritising the need for air renovation within their establishments, and they are therefore failing to minimise the risk of airborne viral transmission between their customers.

## Data Availability

All data produced in the present study are available upon reasonable request to the authors

## Acknowledgements

IDAEA-CSIC is a Centre of Excellence Severo Ochoa (Spanish Ministry of Science and Innovation, Project CEX2018-000794-S).

